# Gait Analysis for Thigh-Worn Accelerometry A Data Processing Pipeline using Data-Driven Approaches

**DOI:** 10.1101/2025.11.18.25339671

**Authors:** Claas Lendt, Martin Grimmer, Ingo Froböse, Tom Stewart

## Abstract

**Introduction:** Thigh-worn accelerometry is becoming increasingly popular in large-scale cohort studies for quantifying movement behaviour. Gait characteristics are associated with various health conditions and can be used to predict fall risk, monitor disease progression and evaluate rehabilitation outcomes. However, accurate gait assessment typically requires controlled laboratory conditions, that may not reflect real-world mobility. In this context, data-driven algorithms and machine learning approaches hold promise for extracting accurate gait parameters from raw accelerometer data.

**Objective:** We developed and evaluated a machine learning-based processing pipeline that uses activity classification to detect walking sequences, estimate walking speed, and identify gait events from raw thigh-worn accelerometer data, enabling accurate assessment of free-living gait.

**Methods:** We integrated an existing activity classification algorithm into the pipeline and evaluated its performance in free-living conditions. Walking speed was estimated based on stride frequency and body height. We then trained a temporal convolutional network model to predict the probability of gait events (i.e. initial and final contact) in healthy adults walking at various speeds on different inclines. All three components of the data processing pipeline were evaluated externally using various independent datasets.

**Results:** The activity classification model achieved F1 scores ≥ 0.95 for walking in both adults and older adults. Walking speed was estimated with a mean absolute percentage error of 11.5%, and with a bias of 0.02 m/s. The gait event detection model demonstrated high accuracy, with a mean recall ≥ 0.94, precision ≥ 0.98, and mean absolute errors of 20 ms and 31 ms for initial and final contacts, respectively.

**Conclusion:** Accurate gait analysis in free-living conditions can be achieved by combining data-driven and machine learning approaches with thigh-worn accelerometer data. The developed pipeline can support the analysis of existing thigh-worn accelerometer datasets and enable continuous gait monitoring outside laboratory settings over several days. However, the developed methods for estimating walking speed and gait events require validation in a more diverse sample and in truly unrestricted free-living conditions.

## Introduction

Thigh-worn accelerometers have become increasingly popular in physical activity research, as they allow researchers to quantify the physical behaviour of individuals under free-living conditions over multiple days. Several large-scale cohort studies within the Prospective Physical Activity, Sitting and Sleep (ProPASS) consortium now use thigh-worn accelerometers [1]. When combined with data-driven algorithms and machine learning approaches, these devices allow the classification of various activity types and postures under free-living conditions [2], [3]. The implementation of novel activity classification algorithms can subsequently enable new scientific evidence of associations between activity types as well as their patterns and health outcomes, which would otherwise not be feasible [4]. Due to the high classification accuracy of activity types and their placement on the lower extremities, thigh-worn accelerometers could potentially be used to measure novel digital biomarkers, such as gait characteristics, by analysing the raw acceleration signal.

Human gait is a sensitive indicator that can provide valuable insights into various health conditions, including neurological disorders, musculoskeletal issues, and the effects of ageing [5], [6]. Changes in gait patterns can reveal important clinical information about health status and disease risk, such as the onset of Parkinson’s disease, post-surgery rehabilitation progress, and the risk of falling [7], [8]. For instance, walking speed differs between fallers and non-fallers, with a slower walking speed being associated with an increased risk of falling [7], [9], [10]. In people with Parkinson’s disease, temporal gait characteristics such as stance time, stride time and stride time variability have been proposed as predictive markers for disease onset and progression [11], [12], [13], [14].

Until recently, accurate gait analysis was mostly confined to laboratory settings and costly measurement methods, such as instrumented treadmills, pressure mats or optical motion tracking systems. However, wearable devices such as accelerometers and inertial measurement units (IMUs), when paired with data-driven algorithms, now offer the opportunity to measure gait unsupervised, in real-life settings and over the course of multiple days. Notably, studies suggest that free-living gait differs significantly from laboratory gait and may be more clinically relevant [6], [15]. Accurate detection of gait events, such as initial contacts (IC; when the foot first contacts the ground) and final contacts (FC; when the foot leaves the ground) is critical to any gait-specific analysis. It enables the computation of spatiotemporal gait variables such as walking speed, stance time and swing time [16], and the segmentation of individual strides to be used for subsequent model input [17], [18]. Recent studies have demonstrated that applying machine learning techniques to IMU data can significantly improve gait event detection [19], [20]. Previous research on free-living gait monitoring has focused primarily on signals from the foot, shank, or lower back, combining both acceleration and gyroscope signals from wearable IMUs [16], [20], [21].

With a projected total sample size of over 110,000 participants, large epidemiological cohort studies, such as HUNT4 [22] as well as AusPASS and ActiveLife (with ongoing data collection), offer a unique opportunity to study risk factors and predictive markers for disease onset based on thigh-worn accelerometry data [23]. To enable research into potential associations between human gait parameters and health outcomes, there is a clear need for a validated, open-source method to perform gait analysis solely based on thigh-worn acceleration data. Building on previous findings regarding the predictive value of gait parameters for various health outcomes and the availability of large-scale datasets, we aim to develop and evaluate a new data processing pipeline for analysing free-living gait based on continuous raw thigh-worn accelerometer data. Our work integrates previous and novel machine learning models to provide a comprehensive, open-source gait analysis pipeline that improves our ability to analyse gait in various clinical and research contexts. We make use of data collected from six different studies to (a) evaluate a previously trained activity classification model in free-living conditions, (b) modify and evaluate an existing walking speed estimation approach and (c) train and evaluate a gait event detection model based on machine learning.

## Methods

We have developed a data processing pipeline (**Figure 1**) that enables gait analysis from continuous, raw three-dimensional thigh acceleration data. The proposed pipeline consists of several key steps:

**Figure 1:**
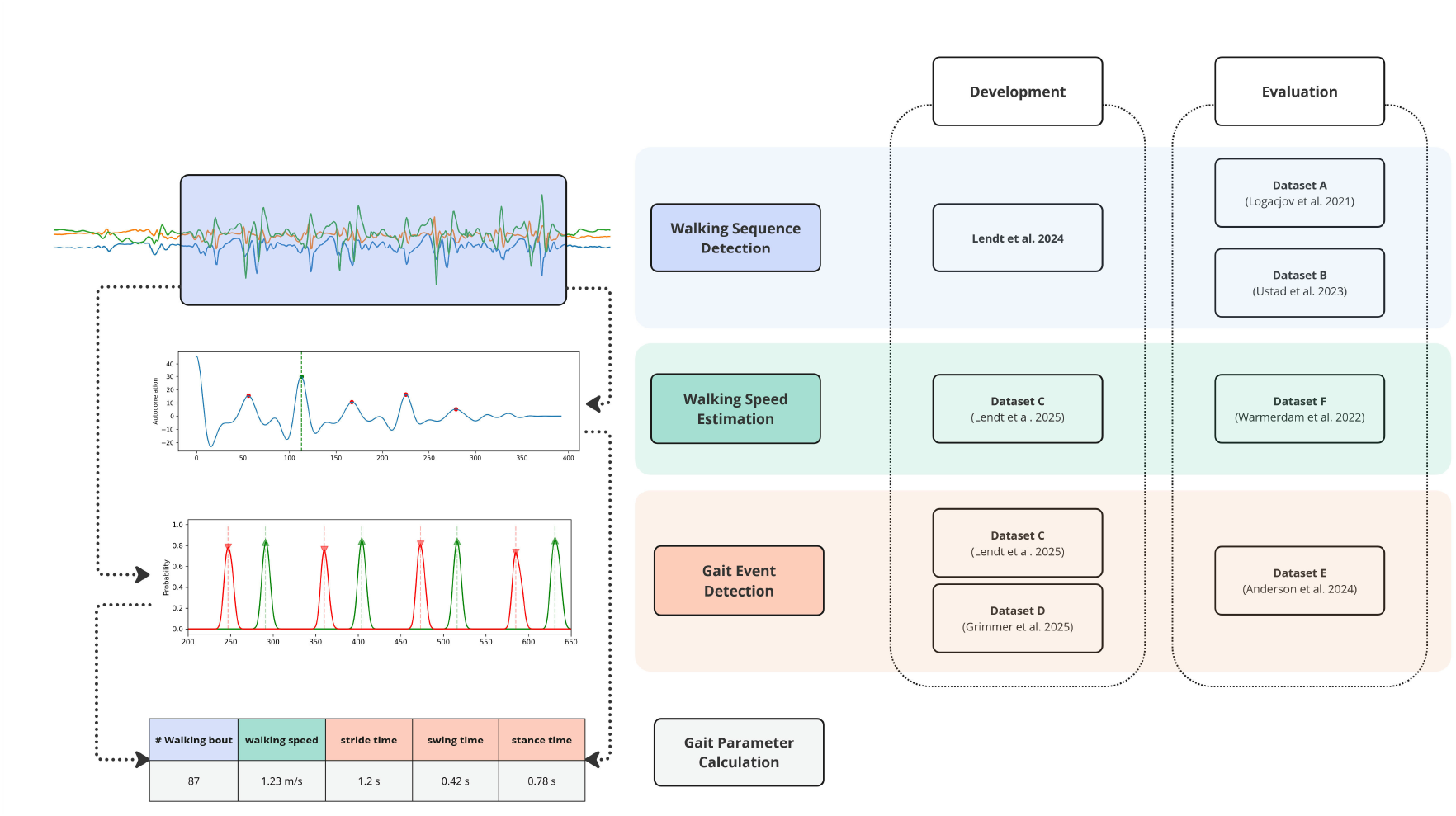
Schematic data processing pipeline with the key steps as well as the respective datasets used for development and evaluation. Raw three-dimensional acceleration data and the individual body height are required as input.

1. .**Walking Sequence Detection**: A machine learning activity classification algorithm identifies walking sequences based on 4-second windows of acceleration data.
2. .**Walking Speed Estimation**: For each 4-second window detected as walking, the walking speed is estimated based on the stride frequency and body height.
3. .**Gait Event Detection**: A machine learning algorithm predicts the probability of initial (IC) and final (FC) contact events within each walking sequence.
4. .**Gait Parameter Calculation**: Bout-level gait parameters are derived from steps 1 to 3, including temporal parameters (e.g. stride time) as well as mean and maximal walking speed.

### Datasets

We used a total of six different datasets to develop and evaluate the data processing pipeline (**Table 1**). The use of different datasets to evaluate different pipeline steps was necessary, as no single dataset was available that comprises gold-standard references for all respective outcomes. Two datasets (A and B) with data from a total of n = 49 participants were used for a free-living evaluation of an existing activity classification algorithm. Dataset C was used to develop the walking speed estimation approach and dataset F with n = 21 participants was used for evaluation. Two of the datasets (C and D) with a total of n = 34 participants were used for the development of a gait event detection algorithm. Dataset E with n = 59 participants was used to evaluate the gait event detection algorithm. All six underlying studies were approved by the respective ethics committees and in accordance with the Declaration of Helsinki. For each study, participants gave written informed consent prior to data collection. More details on the individual datasets and data processing can be found in Supplementary File 1.

**Table 1:** Characteristics of the individual datasets used for development and evaluation of the data processing pipeline.

| Dataset | A | B | C | D | E | F |
| --- | --- | --- | --- | --- | --- | --- |
| **Data source** | Logacjov et al. 2021 [24] | Ustad et al. 2023 [25] | Lendt et al. 2025 [26] | Grimmer et al. 2019 [27] | Anderson et al. 2024 [28] | Warmerdam et al. 2022 [29] |
| **Sample size** | 31 | 18 | 21 | 13 | 59 | 21 |
| **Participant characteristics** | Female = 8 (36%)<br>Age = $38.6 \pm 14$ yrs<br>BMI = $23.1 \pm 2.3$ kg/m<sup>2</sup> | Female = 9 (50%)<br>Age = $79.6 \pm 8$ yrs<br>BMI = $26.8 \pm 2.7$ kg/m<sup>2</sup> | Female = 9 (43%)<br>Age = $26.7 \pm 4$ yrs<br>BMI = $23.4 \pm 1.6$ kg/m<sup>2</sup> | Female = 5 (38%)<br>Age = $26 \pm 4$ yrs<br>BMI = $21.8 \pm 2.3$ kg/m<sup>2</sup> | Female = 19 (32%)<br>Age = $67.9 \pm 7.6$ yrs<br>BMI = $26.4 \pm 4.4$ kg/m<sup>2</sup> | Female = 8 (38%)<br>Age = $34.6 \pm 14$ yrs<br>BMI = $23.3 \pm 3.2$ kg/m<sup>2</sup> |
| **Context** | Free-living; unstructured protocol | Free-living; semi-structured protocol | Standardised free-living and laboratory | Laboratory | Laboratory; self-paced walking through a building | Laboratory |
| **Activities included** | Sitting, lying, walking, running, standing, stairclimbing, cycling | Sitting, lying, walking, standing, stairclimbing | Treadmill walking; outdoor walking (flat) | Treadmill walking (flat and incline) | Indoor walking and stairclimbing | Walking (over ground) at slow, preferred and fast speed |
| **Reference measure** | Wearable chest-mounted video camera | Wearable chest-mounted video camera | Pressure-sensor insoles | Instrumented treadmill | Pressure-sensor insole | Optical motion capture |
| **Sensor type** | Axivity AX3 | Axivity AX3 | Axivity AX6 | SimpleLink SensorTag | Xsens MTw Awinda | Noraxon myoMOTION |
| **Accelerometer sampling frequency** | 50 Hz | 50 Hz | 100 Hz | 133 Hz | 100 Hz | 200 Hz |

### Pre-processing

Each dataset contained three-dimensional thigh acceleration data sampled at frequencies ranging from 50 to 200 Hz. To ensure consistency across varying datasets, we implemented a standardized preprocessing workflow with three main steps. First, raw acceleration signals were filtered using an eighth-order Butterworth low-pass filter with a 20 Hz cutoff frequency to remove high-frequency noise while preserving the relevant movement signals. Second, when necessary, data were resampled to a uniform 100 Hz sampling rate using the actipy Python package [30]. Third, we standardized the accelerometer axis orientations with the X-axis representing the cranial-caudal direction, the Y-axis representing the medial-lateral direction, and the Z-axis representing the anterior-posterior direction, all defined relative to a quiet standing posture.

### Walking Sequence Detection

The gait analysis pipeline integrates a previously developed activity classification model – based on a hybrid Convolutional Neural Network (CNN) and Bidirectional Long-Short Term Memory (BiLSTM) model architecture [18]. The CNN-BiLSTM model classifies 4-second windows as either sedentary (sitting or lying), standing, walking, running or cycling. The model was trained on a pooled dataset comprising both laboratory and free-living activity data from three different datasets (n = 69 participants) and is openly available on GitHub (https://github.com/claaslendt/ catse3). Datasets A and B were used to evaluate the classification accuracy of the CNN-BiLSTM activity classification model in free-living conditions. The combined dataset comprises different free-living physical activities, including walking, stairclimbing, running, standing, sitting, reclining, lying down, and cycling. Reference labels were obtained from video annotations. For each dataset, non-overlapping 4-second windows were created across each individual activity sequence. Only windows with a single true label were considered, meaning that no ambiguous windows comprising two or more activities were used. Each of the finally extracted 4-second windows included 400 samples of continuous three-dimensional acceleration data and a single reference activity label.

### Walking Speed Estimation

A previously published method for estimating walking speed was modified and integrated into the analysis pipeline. The approach exploits the relationship between walking speed and stride frequency using the following equation:

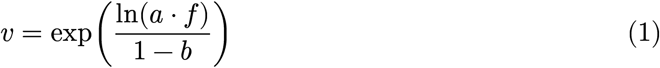

where v is the walking speed in meters per second and f is the stride frequency in strides per second, normalized by the leg length [31]. In the original study, the parameters a and b were individually identified per participant using a standardised calibration procedure. We calculated the average parameter values across all participants from the original study and set a = 2.064 and b = 0.349. Further, leg length is estimated from the participants height scaled by a factor of 0.53 [32]. The stride frequency is estimated within each 4-second walking sequence based on the acceleration in the anterior-posterior axis. The acceleration signal is low-pass filtered using a fourth-order Butterworth filter with a cut-off frequency of 5 Hz. The stride time is then estimated as the lag associated with the largest peak in the autocorrelation between lags of 0.5 and 3 seconds (**Figure 2**). These lower and upper thresholds are implemented to minimise the risk for detecting false positive peaks outside the anticipated range of human stride times [33]. Finally, stride frequency (in Hz) is calculated from the estimated stride time (in seconds). Treadmill walking data in dataset C was used to develop the autocorrelation approach. Overground walking data in dataset F was used to evaluate the walking speed estimations against the reference walking speed estimated from optical motion capture.

**Figure 2:**
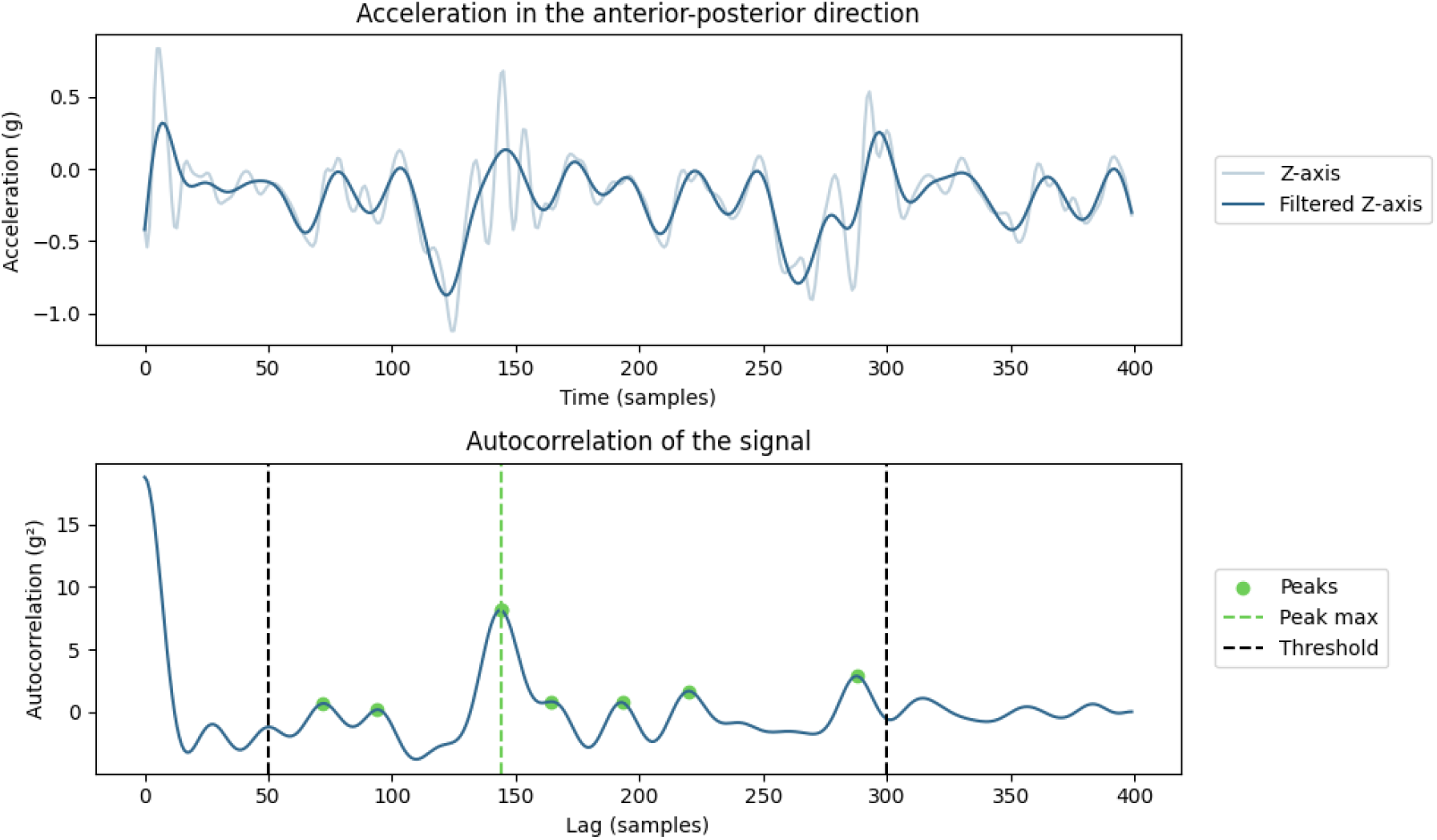
Example of the raw and low-pass filtered anterior-posterior acceleration (Z-axis) of a 4-second walking sequence (top) and the corresponding autocorrelation (bottom). Stride time is estimated based on the lag associated with the highest peak of the autocorrelation coefficient between a lag of 0.5 and 3 seconds (dashed green line).

### Gait Event Detection

Datasets C and D were used to develop and train the gait event detection algorithm. These datasets consisted of gait data from 21 and 13 healthy adult participants, respectively. The combined dataset comprised treadmill walking at speeds ranging from 0.5 m/s to 2.1 m/s, both on a level surface and on an inclined surface at angles of ±5 degree and ±10 degree. Additionally, outdoor walking at self-selected speeds was performed. A non-causal temporal convolutional network (TCN) [34] with a fully connected output layer and Softmax activation function was trained to predict the probability of initial contact (IC) and final contact (FC) events for each individual acceleration sample in a walking sequence. Further details on the architecture, training and optimisation of the TCN model can be found in Supplementary File 2. An additional filtering approach was designed to minimise false positive predictions and applied separately to the IC and FC predictions. The resulting probabilities were first smoothed using a Gaussian filter with three standard deviations. Peaks with a gait event probability greater than 0.4 and a minimum interpeak distance of 0.5 s were then identified. Both values were optimised using an iterative approach and visual inspection of the predictions during model development. The resulting peaks were then considered to be the predicted gait events. After training the model on the combined training and validation set, we used data from Dataset E to evaluate the performance and accuracy of the final model. The dataset contains indoor walking data from n = 21 healthy adult participants as well as n = 38 individuals with Parkinson’s disease who each completed a standardised self-paced walking course through a building, including level walking and stairclimbing.

### Gait Parameter Calculation

Temporal gait parameters were calculated based on predicted gait events, with implausible values removed through filtering. Stride times were calculated as the time between two consecutive IC events. These were filtered to remove values above 3 s and below 0.5 s, as well as strides covering two FC events, which indicated a missing IC event. Stance times were calculated as the time between an IC event and the subsequent FC event. These were filtered to remove times greater than 2 s or smaller than 0.2 s. Swing times were calculated as the time between an FC event and the subsequent IC event. Swing times greater than 1 s or smaller than 0.1 s were removed.

## Statistical Analysis

The metrics used for the evaluation of the walking sequence detection model included recall, precision, and F1 score. Recall (or sensitivity) was calculated as the proportion of all true walking sequences that were identified by the model. Precision was calculated as the proportion of predicted walking sequences that were true walking sequences. The F1 score was calculated as the harmonic mean of recall and precision. For the walking speed estimation, we calculated the mean difference (bias), the root mean squared error (RMSE) and mean absolute percentage error (MAPE) from the reference and estimated walking speed of each individual trial. For the gait event detection, predicted gait events were matched with the corresponding reference gait events based on the closest reference event. Predicted gait events were considered true positive events when they were within a 0.5 s tolerance window centred around a reference gait event [19], [21]. Predicted gait events not within this time window were considered false positive events. Overall detection performance metrics included recall, precision and F1 score. We performed a true positive evaluation to estimate the time error of the gait event predictions. The time difference between each predicted true positive gait event and corresponding reference gait event was calculated. The gait-specific parameters stride time, stance time and swing time were calculated based on all reference gait events and predicted gait events. Mean absolute errors (MAE) for the gait-specific parameters were calculated per walking sequence by comparing the reference method with the proposed pipeline.

## Results

### Walking Sequence Detection

A total of 38,364 (Dataset A) and 10,285 (Dataset B) 4-second activity sequences were evaluated (**Figure 3**). When the CNN-BiLSTM activity classification model was evaluated on Dataset A, it showed an unweighted mean precision of 0.94, mean recall of 0.96 and mean F1 score of 0.95 across all five activity classes. For the walking class, recall, precision as well as F1 score were 0.95. For Dataset B, the classification model showed an unweighted mean precision of 0.99, mean recall of 0.98 and F1 score of 0.98. For walking, precision was 0.99, recall was 0.98 and F1 score was 0.99.

**Figure 3:**
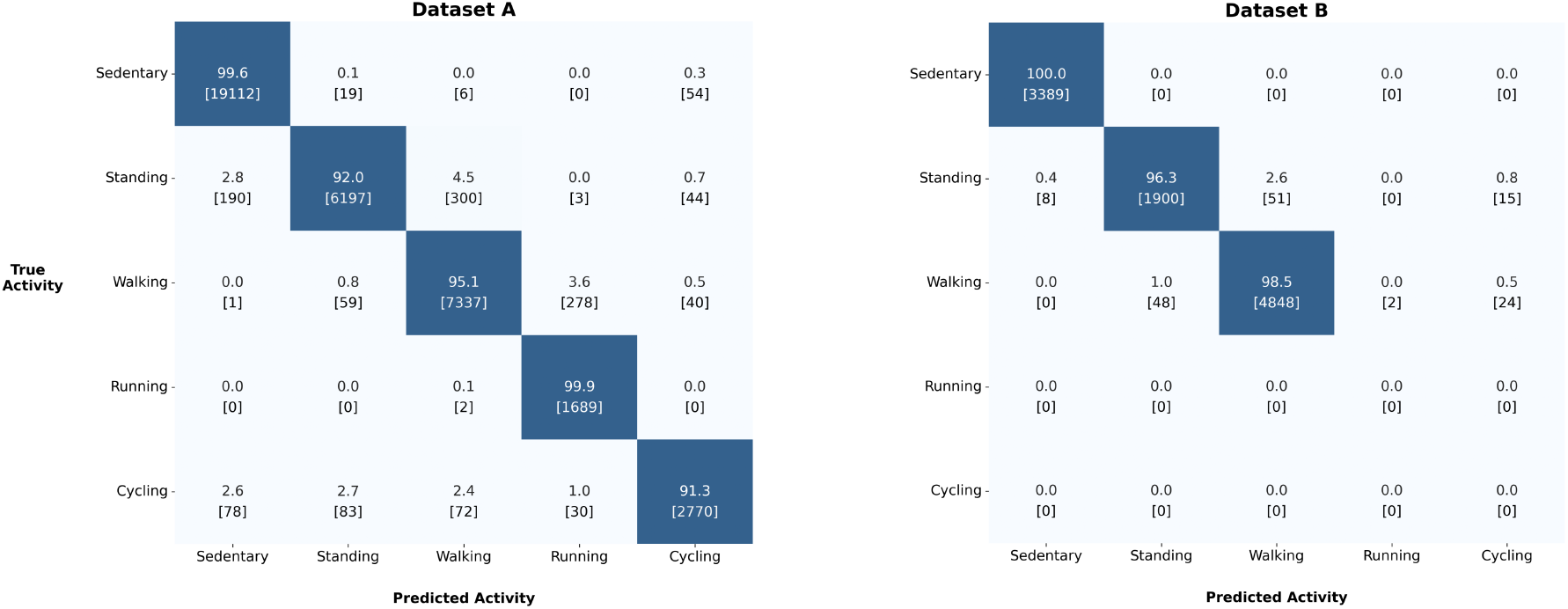
Confusion matrices for the CNN-BiLSTM activity classification model [18] when evaluated on Dataset A (left) and Dataset B (right). Percentages [samples] are calculated row-wise.

### Walking Speed Estimation

Across all three self-paced walking speed conditions in Dataset F, walking speed ranged from 0.46 to 2.61 m/s (see **Figure 4**). The overall RMSE was 0.18 m/s, the MAPE was 11.5%, and the bias was0.02 m/s. For the preferred walking speed condition, the reference walking speed ranged from 0.89 to 1.85 m/s, with an associated RMSE and MAPE of 0.12 m/s and 7.1%, respectively.

**Figure 4:**
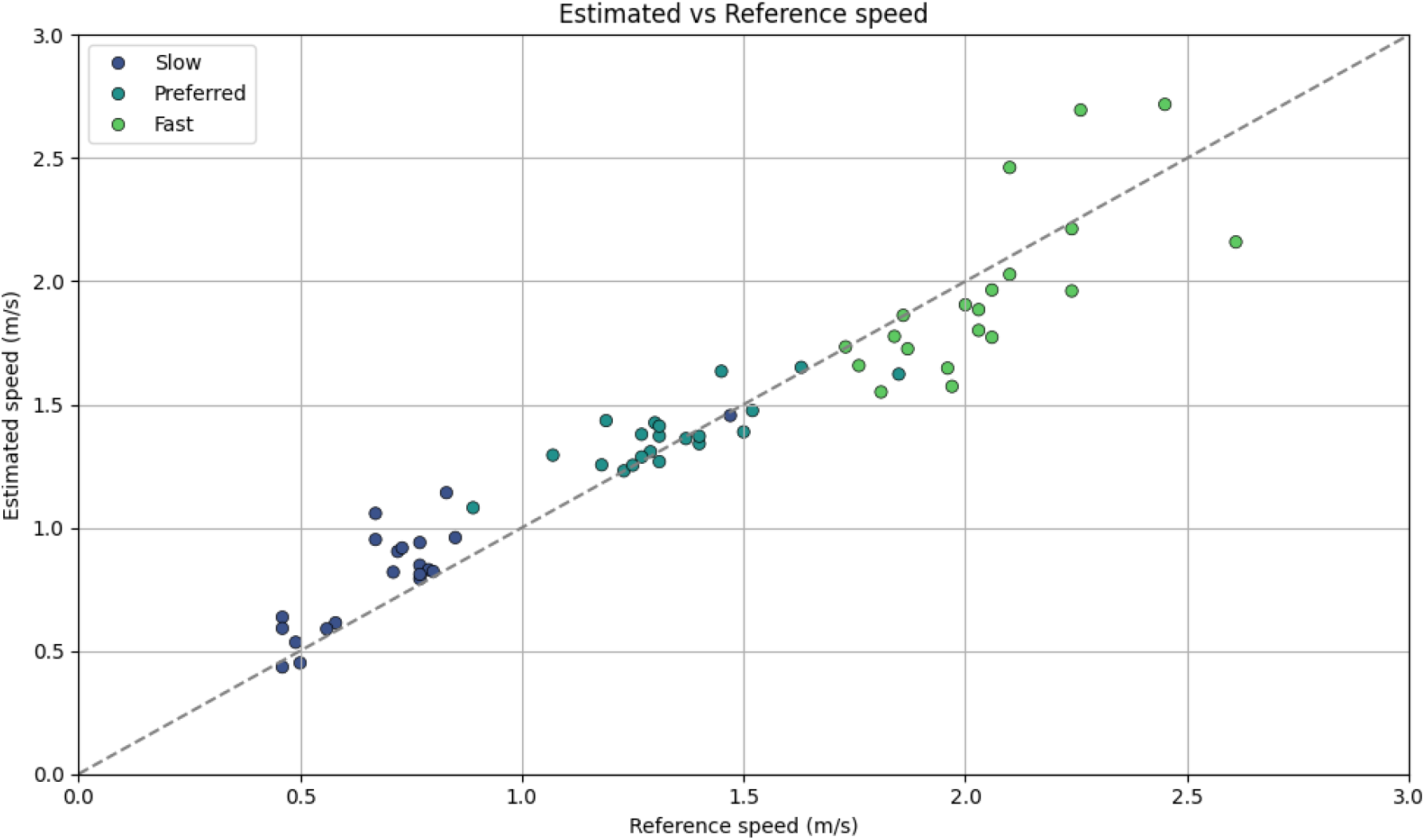
Scatter plot of estimated walking speed against reference walking speed for all trials in Dataset F. The dashed line represents the line of identity for reference.

### Gait Event Detection

Dataset E contained a total of 9,174 gait events (**Table 2**). The TCN gait event detection model achieved mean recall values of 0.97 (95% CI: 0.97, 0.98) for IC events and 0.94 (95% CI: 0.94, 0.95) for FC events across all participants. The mean F1 scores were 0.98 (95% CI: 0.98, 0.99) for IC events and 0.96 (95% CI: 0.96, 0.97) for FC events. The predicted IC and FC events as well stride time, stance time and swing time showed mean time differences (bias) within ±20 ms (**Figure 5**). The MAE for IC and FC events were 20 ms and 31 ms, respectively. The MAE for the spatiotemporal gait parameters were 15 ms (stride time), 23 ms (stance time) and 22 ms (swing time).

**Table 2:** Gait event detection performance metrics for initial contact (IC) and final contact (FC) events when evaluated on Dataset E.

|  | Initial Contact (IC) | Final Contact (FC) |
| --- | --- | --- |
| Reference Events | 4,590 | 4,584 |
| Predicted Events | 4,486 | 4,395 |
| True Positives | 4,455 | 4,316 |
| False Positives | 31 | 79 |
| False Negative | 135 | 268 |
| Recall (mean [95% CI]) | 0.97 [0.97, 0.98] | 0.94 [0.94, 0.95] |
| Precision (mean [95% CI]) | 0.99 [0.99, 1.0] | 0.98 [0.98, 0.99] |
| F<sub>1</sub> score (mean [95% CI]) | 0.98 [0.98, 0.99] | 0.96 [0.96, 0.97] |

**Figure 5:**
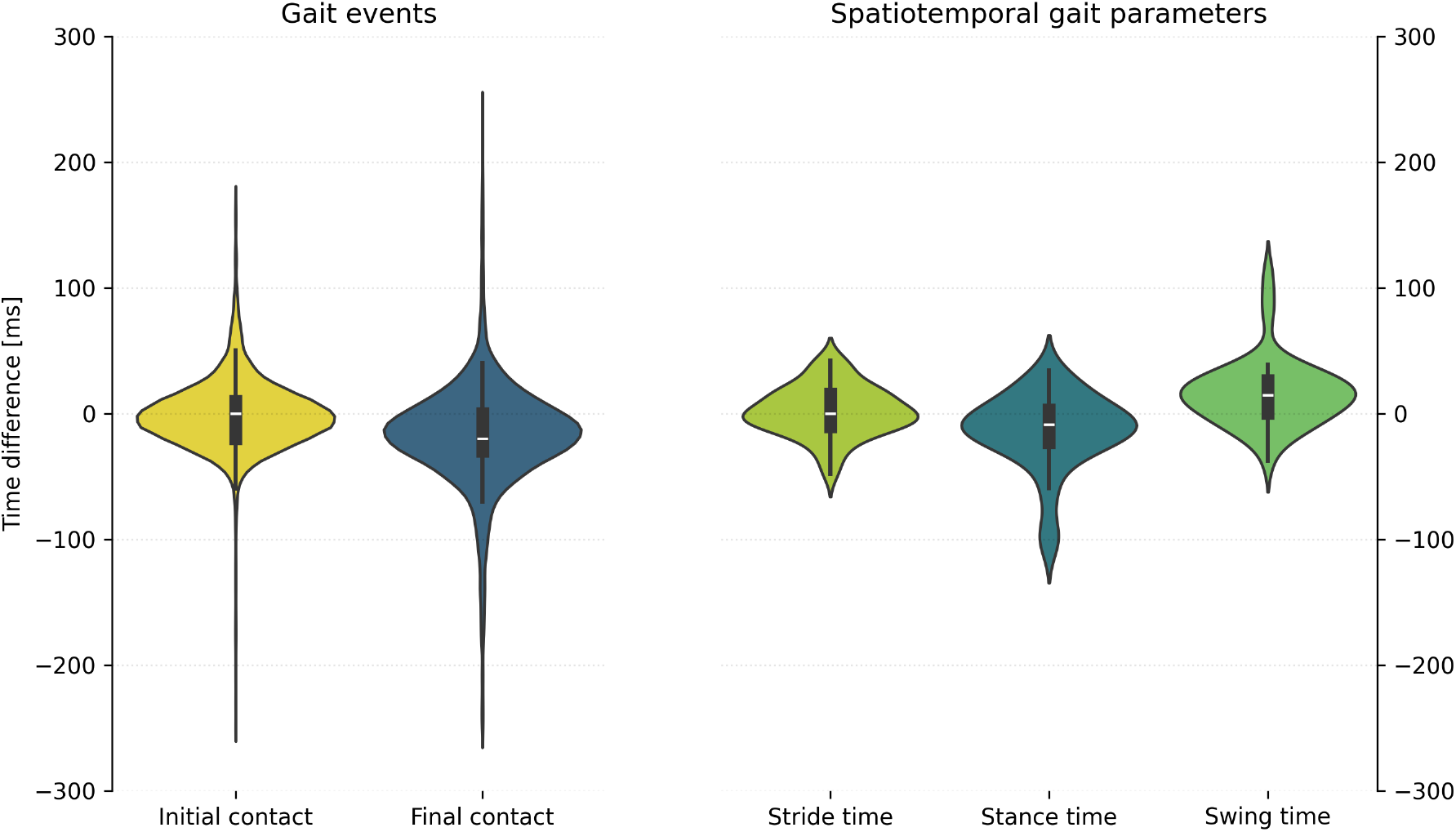
Time differences for the individual predicted and reference gait events for the true positive evaluation (left). Bout-wise time differences between predicted and reference spatiotemporal gait parameters (right).

## Discussion

Human gait is associated with various health outcomes and useful as a predictive biomarker. A validated data processing and gait analysis pipeline specifically for thigh-worn accelerometers would enable the research community to analyse gait-related outcomes using already existing, large-scale datasets. In this study, our aim was to develop and evaluate a novel gait analysis pipeline based on supervised machine learning and data-driven approaches.

When compared with gold-standard video observations, the walking sequence detection model achieved an F1 score of at least 0.95 for walking in free-living conditions in both adults and older adults. The activity classification model’s classification performance is slightly superior to that of previous works using decision tree-based machine learning approaches, such as extreme gradient boosting [2], [25], [35]. Walking sequence detection can be considered highly accurate and sufficient to capture most walking sequences, enabling meaningful gait analysis representative of an individual’s gait.

Walking speed was estimated with a MAPE of 11.5% and minor positive bias of 0.02 m/s. Considering the laboratory condition and apparently healthy individuals, this error is slightly larger when compared to a previously, extensively validated gait analysis pipeline using data from a single IMU placed on the lower back, reporting a MAPE of 10.3% [16]. The approach chosen for the proposed pipeline, which is based on the relatively simple relationship between normalised stride frequency and walking speed, resulted in an overall RMSE of 0.18 m/s. Importantly, the estimation error varied by walking speed: the error was lower during slow (RSME: 0.16 m/s) and preferred walking speed (0.12 m/s), but considerably higher during fast walking speeds above 1.8 m/s (0.25 m/s). Our results are consistent with a previous study that trained a support vector machine regression model for thigh-worn accelerometers based on 32 time– and frequency-domain features in 5-second windows, achieving an RMSE of 0.15 m/s with walking speeds up to 1.7 m/s [36]. Notably, preferred and fast outdoor walking speeds have been shown to be within 1.2 to 1.8 m/s, suggesting that the lower precision at walking speeds above 1.8 m/s may not be of concern for most individuals [37], [38]. Overall, the error margin for walking speed estimations falls within an acceptable range for estimating walking speed on a group-level in larger-scale cohort. However, for clinical adult populations, a minimal clinically important difference for walking speed between 0.10-0.20 m/s has been reported [39], [40]. Given our results and the evaluation in a healthy adult population, the walking speed estimations may not yet be sufficiently accurate for use in clinical settings.

A TCN deep learning architecture was used to identify gait events, achieving high F1 scores of 0.98 (IC) and 0.96 (FC), with a low bias of less than 20 ms. We found a high level of agreement for the timing of gait events and the derived spatiotemporal gait parameters. The MAE for the temporal gait parameters stance time (23 ms) and swing time (22 ms) are comparable to those of two previous studies that used a similar deep learning approach based on IMU data from the shank or foot [19], [20]. The authors report MAE values of 30 ms for both stance time and swing time, when evaluated in healthy adults under free-living conditions. Furthermore, the agreement of the estimated spatiotemporal gait parameters, specifically stance and swing time, is comparable to that of a previous study using thigh acceleration and a deterministic filtering approach [41]. However, it should be noted that this previous study only evaluated treadmill walking, which likely resulted in an overestimation of accuracy compared to free-living walking conditions.

### Strengths and Limitations

The sensors, settings and placements used in the various datasets are common in physical activity research, making our results broadly applicable to other studies and datasets. For the development and evaluation of our proposed pipeline, we used gait data from various available datasets that comprise a high level of real-world walking conditions. We successfully evaluated the different modules of the pipeline using independent, high-quality datasets with gold-standard criterion measures for each outcome. This allowed us to further validate the previously developed CNN-BiLSTM activity classification approach using two datasets comprising free-living activity data from adults and older adults, with video-annotated activity labels used as a reference. However, we were unable to compare the walking speed estimates against an appropriate reference under free-living conditions. Despite this important limitation, we were able to evaluate the accuracy across a wide range of walking speeds, from 0.46 to 2.61 m/s, in a controlled laboratory setting using a gold-standard reference measure.

Although several studies have developed algorithms for gait event detection, these are often only evaluated using data collected under standardised laboratory conditions, such as walking on a treadmill or over a short distance in a straight line. Therefore, it is unclear how accurate these algorithms are in real-life situations. To address this issue, we trained our TCN model using two datasets comprising indoor and outdoor walking at various speeds and inclines. We were also able to evaluate the model’s performance against a gold standard reference measure under semi-standardised conditions that closely resemble real life (i.e. an indoor walking course through a building), thereby increasing the generalisability of its performance. However, the model’s ability to accurately predict gait events may be affected by musculoskeletal or neurological diseases and ageing, since these factors can impact individual gait patterns. Therefore, additional validation of our results is necessary, including testing on a more heterogeneous sample comprising different age groups and health conditions.

Importantly, accessible models and processing approaches remain an ongoing challenge in the field of physical activity research [42]. The complete pipeline, which was developed as part of this study, is available as open-source software on the GitHub repository (https://github.com/claaslendt/twaga). This enables the research community to use, evaluate and build upon it.

### Future investigations

While the study produced favourable results, there are several areas that need to be addressed in future work. Firstly, it would be beneficial to establish convergent validity using a large-scale existing dataset, such as the HUNT4 study [22]. If gait characteristics are found to be associated with specific health outcomes in the expected direction (e.g. a decline in walking speed being associated with a decline in self-rated health), this would strengthen our understanding of the robustness and applicability of the pipeline at a group level. Secondly, the accuracy of walking speed estimation could be improved by exploring machine learning-based approaches that use body height and acceleration as inputs, as opposed to acceleration alone [36]. Furthermore, step length and walking speed could potentially be estimated based on segmented strides instead of generic 4-second input windows, providing more precise estimates [43]. Thirdly, gait characteristics change depending on the walking surface, i.e. whether it is flat or inclined. Therefore, the activity classification algorithm could be extended to distinguish between walking, incline walking and stair climbing. This could enable walking-type specific gait analysis and provide new insights, such as using stair-climbing speed as an indicator of aerobic capacity. Lastly, the current pipeline implementation only covers the calculation of basic bout-average metrics, such as stride time, swing time and stance time, as well as walking speed. Future versions of the pipeline could incorporate additional gait-related variables, such as variability metrics or average stride acceleration profiles [8].

## Conclusion

The developed data-driven gait analysis pipeline for thigh-worn accelerometers demonstrated high accuracy in detecting walking sequences, estimating walking speed, and identifying gait events, when tested using independent datasets. Overall, the pipeline is an important step towards creating a validated, open-source tool for analysing gait in existing accelerometer datasets. Future work should extend the validation to clinical populations, explore more advanced machine learning approaches for estimating walking speed and incorporate additional gait characteristics to enable a more comprehensive mobility assessment.

## Acknowledgements

We would like to thank all participants taking part in any of the underlying studies for their time and commitment. We further thank all authors and involved research teams of the underlying studies for sharing their data.

## Data Availability

Datasets A and B are openly accessible and can be downloaded from the GitHub repository (https://github.com/ntnu-ai-lab/harth-ml-experiments). Dataset C is openly accessible and can be downloaded from the Zenodo repository (https://doi.org/10.5281/zenodo.17629130). Dataset D is not openly accessible but may be shared upon reasonable request to the corresponding author of the underlying study. Dataset E can be accessed and downloaded after registration from the Synapse repository (https://www.synapse.org/Synapse:syn52540892/files). Dataset F is openly accessible and can be downloaded from the Figshare repository (https://doi.org/10.6084/m9.figshare.20238006.v2).

