## Supplementary File 1 for "Gait Analysis for Thigh-Worn Accelerometry A Data Processing Pipeline using Data-Driven Approaches"

### Datasets used for the development and evaluation of the pipeline

#### Dataset A: HARTH (Logacjov et al. 2021)

The HARTH dataset contains free-living activity data from n = 31 healthy adults, who had an accelerometer (Axivity AX3; Axivity Ltd., United Kingdom) placed on the right frontal thigh (10cm above the patella). Participants performed everyday activities with minimal instructions (i.e. to include at least a few minutes of each activity) with a subset focussed on performing walking, running and cycling. Acceleration data was annotated based on synchronized videos from a wearable chest-worn video camera (GoPro Hero3+; GoPro Inc., USA). Twelve activities and postures were annotated. The underlying study was approved by the Mid-Norway Regional Committee for Ethics in Medical Research (ref. 2015/1432). Additional information regarding the data collection can be found in the original publication:

Logacjov, A., Bach, K., Kongsvold, A., Bårdstu, H. B., & Mork, P. J. (2021). HARTH: A Human Activity Recognition Dataset for Machine Learning. Sensors, 21(23), 7853. https://doi.org/10.3390/s21237853

For our analysis, we collapsed these labels into five basic activity types (Table S1). We sampled non-overlapping 4-second windows and only included windows with a single true label, meaning that no ambiguous 4-second windows comprising two or more activities were used.

**Table S1**: Annotated and processed activity labels in the HARTH dataset.

| Processed labels | Annotated labels |
|---|---|
| sedentary | sitting, lying |
| standing | standing, shuffling |
| walking | walking, stairclimbing (up/down) |
| running | running |
| cycling | cycling (sit / stand / sit,inactive / stand,inactive) |

### Dataset B: HAR70+ (Ustad et al. 2023)

The HAR70+ dataset contains free-living activity data from n = 18 older adults aged 70 years or older, who had an accelerometer (Axivity AX3; Axivity Ltd., United Kingdom) placed on the right frontal thigh (10cm above the patella). Participants performed a semi-structured free-living protocol including lying, sitting, standing and walking within their home as well as outdoors. Five participants used walking aids during data collection. Acceleration data was annotated based on synchronized videos from a wearable chest-worn video camera (GoPro Hero 8; GoPro Inc., USA). Seven activities and postures were annotated. The underlying study was approved by the Norwegian Centre for Research Data (ref. 515701). Additional information regarding the data collection can be found in the original publication:

Ustad, A., Logacjov, A., Trollebø, S. Ø., Thingstad, P., Vereijken, B., Bach, K., & Maroni, N. S. (2023). Validation of an Activity Type Recognition Model Classifying Daily Physical Behavior in Older Adults: The HAR70+ Model. Sensors, 23(5), 2368. https://doi.org/10.3390/s23052368

For our analysis, we collapsed these labels into three basic activity types (Table S2). We sampled non-overlapping 4-second windows and only included windows with a single true label, meaning that no ambiguous 4-second windows comprising two or more activities were used.

**Table S2**: Annotated and processed activity labels in the HAR70+ dataset.

| Processed labels | Annotated labels |
|---|---|
| sedentary | sitting, lying |
| standing | standing, shuffling |
| walking | walking, stairclimbing (up/down) |

### Dataset C: Lendt et al. (2025)

The dataset contains laboratory and outdoor gait data from n = 21 healthy adult participants. Participants completed four two-minute walking trials on a treadmill at speeds ranging from 2 to 5 km/h. They also completed a five-minute walk outdoors at their preferred speed on level ground. Vertical ground reaction force (GRF) was measured using force-sensing insoles (loadsol pro, novel GmbH, Germany). Thigh acceleration was measured using an IMU (Axivity AX6; Axivity Ltd., United Kingdom) placed on the lateral side of the dominant leg, midway between the hip and knee. The IMU was attached directly to the skin using medical adhesive tape. Insole force data and IMU data were sampled at 100 Hz. The underlying study was approved by the German Sport University Ethics committee (ref. 107/2023).

To synchronise the insole and IMU signals, participants performed a series of standardised heel drops, lifting their heels off the ground and dropping them rapidly back down again with straight legs. The peaks in both signals were subsequently used to align the two timeseries.

The initial contacts (IC) and final contacts (FC) during walking were identified from the GRF using an adaptive threshold of 25 N + 10th percentile of the GRF during the measurement to account for trials with zeroing errors of the insole [1]. The identified gait events were verified by checking for IC or FC events within 0.5 seconds of each other, as well as for alternating IC and FC events within the data. The GRF and identified gait events were then visually inspected and manually corrected where necessary.

### Dataset D: Grimmer et al. (2019)

The dataset comprises treadmill gait data from n = 13 healthy adults who walked on an instrumented treadmill with speeds ranging between 0.5 and 2.1 m/s. Participants performed level walking as well as inclined and declined walking at ±5° and ±10°. Participants walked continuously for 1 minute for each walking speed and inclination setting. IMUs (SimpleLink SensorTag CC2650STK; Texas Instruments Inc., USA) were placed on the participants frontal thigh using velcro straps. Vertical ground reaction force (GRF) was recorded using the instrumented treadmill (GRAIL, Motek Medical B.V., Netherlands) and subsequently used to identify initial contacts and final contacts. IMUs and the instrumented treadmill were synchronized before the data collection. The underlying study was approved by the Institutional Review Board ETH Zurich. Additional information regarding the data collection can be found in the original publication:

Grimmer, M., Schmidt, K., Duarte, J. E., Neuner, L., Koginov, G., & Riener, R. (2019). Stance and Swing Detection Based on the Angular Velocity of Lower Limb Segments During Walking. Frontiers in Neurorobotics, 13, 57. https://doi.org/10.3389/fnbot.2019.00057

For our analysis, we used the previously identified IC and FC events. We down sampled the data from 1000 Hz to 100 Hz and corrected the axis orientation.

### Dataset E: WearGait-PD (Anderson et al. 2024)

The WearGait-PD dataset contains gait data from approximately 150 individuals with Parkinson's disease, as well as age-matched controls. Participants performed a series of specific walking tasks at both a self-selected pace and a hurried pace over a pressure walkway. A subset of n = 59 participants also performed a free walking task in which they walked along a defined route within the respective research facility, including corridors, corners, and a staircase. All participants wore a full set of 12 IMUs (Xsens MTw Awinda; Movella Inc., USA) and synchronised pressure sensor insoles (Moticon OpenGo; Moticon ReGo AG, Germany). The thigh IMUs were placed laterally, midway between the hip and knee. The IMUs and sensor insoles recorded data at 100 Hz. This study was approved by the Johns Hopkins School of Medicine Institutional Review Board (ref. IRB00234370) and the VA Institutional Review Board (ref. IRB01702255). Additional information regarding the data collection can be found in the original publication:

Anderson, A. J., Eguren, D., Gonzalez, M. A., Khan, N., Watkinson, S., Caiola, M., Hirczy, S. S., Zabetian, C. P., Mills, K., Moukheiber, E., Moro-Velazquez, L., Dehak, N., Motely, C., Muir, B. C., Butala, A., & Kontson, K. (2024). WearGait-PD: An Open-Access Wearables Dataset for Gait in Parkinson's Disease and Age-Matched Controls. Neurology. https://doi.org/10.1101/2024.09.11.24313476

From the available 59 participants in the data, we excluded 2 participants due to missing insole data and one participant due to synchronisation issues which could not be resolved. Additionally, we excluded participants using an assistive device (2) and participants receiving deep brain stimulation (7). The IMUs and sensor insoles were synchronised using a custom hardware configuration, and site-specific corrections were subsequently applied to the data after collection to correct time delays in the sensor insole signal, before the dataset was published. During visual inspection of the data, we noticed minor time shifts between the sensor insoles and the IMU data. We identified the time shift of the sensor insoles by plotting the pressure data from the insoles against the walkway pressure data, shifting the time series of the insoles iteratively until both were aligned (see Figure S1).

Missing IMU acceleration and insole force data were imputed using linear interpolation. The initial contacts and final contacts during walking were identified from the sensor insole GRF data using an adaptive threshold of 50 N + 10th percentile of the GRF during the measurement to account for trials with zeroing errors of the insole.

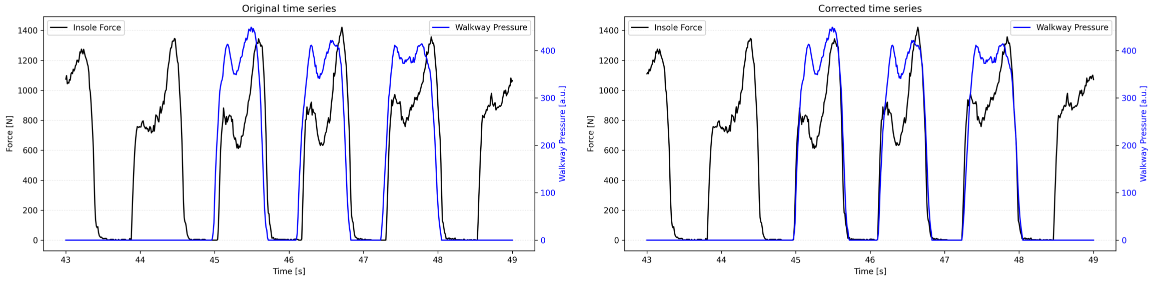


**Figure S1**. Example of misaligned sensor insole data with the original (left) and corrected sensor insole time series (right).

### Dataset F: Warmerdam et al. (2022)

The dataset comprises gait data from n = 21 healthy adults who performed various tasks, including several gait-related activities and everyday tasks, in a laboratory setting. As part of the gait tasks, participants walked along a walkway at self-selected slow, preferred, and fast walking speeds. A 5 m corridor was marked along the walkway. All participants wore a full set of 15 IMUs (myoMOTION, Noraxon Inc., USA). The thigh IMUs were placed laterally, midway between the hip and knee, and secured with elastic straps. Participants were also tracked using an optical motion capture system (Qualisys AB, Sweden) comprising 12 cameras and a set of 47 markers. Both the IMU and motion capture data were sampled at 200 Hz and synchronised using a TTL signal. This study was approved by the Ethics Committee at Kiel University (ref. D438/18, 08/05/2018). Additional information regarding the data collection can be found in the original publication:

Warmerdam, E., Hansen, C., Romijnders, R., Hobert, M. A., Welzel, J., & Maetzler, W. (2022). Full-Body Mobility Data to Validate Inertial Measurement Unit Algorithms in Healthy and Neurological Cohorts. Data, 7(10), 136. https://doi.org/10.3390/data7100136

For our analysis, we down-sampled the data to 100 Hz and used the sternum marker cluster to calculate the distance covered between two annotated timestamps (when first entering the corridor and last exiting the corridor). Walking speed was calculated using the distance and time between both timestamps and used as the reference.
