## Supplementary File 2 for "Gait Analysis for Thigh-Worn Accelerometry A Data Processing Pipeline using Data-Driven Approaches"

### Supplementary File 2
#### Training of the temporal convolutional network (TCN) model

All participant data from datasets C and D were randomly divided at the participant level into two independent datasets, namely a training set (80%) and a validation set (20%). Acceleration data was split into 5-second windows with a 50% overlap, centered around the mean of each axis, and augmented using random rotations and flipping of the axis. The resulting training set consisted of 25,455 and the validation set of 6,711 5-second windows of triaxial acceleration and labelled gait events.

A non-causal temporal convolutional network (TCN) [1] was trained to predict the occurrence of initial contact (IC) and final contact (FC) events within a time series of 3D acceleration data. The TCN architecture included a fully connected layer with a Softmax activation function as the final prediction layer. For the model training, we used weighted categorical cross-entropy as the loss function, a fixed learning rate of 0.0001 and a batch size of 16. The model was trained using Keras (v3.8.0) [2], KerasTuner (v1.4.7) [3] and KerasTCN (v3.5.6) [4].

We performed a grid search to identify the optimal set of hyperparameters for the TCN architecture (**Table S3**). Hyperparameter optimisation was performed by training a model for each set of hyperparameters on the training set and evaluating it on the validation set. The hyperparameter set with the lowest loss was chosen. The final model architecture was defined with 32 filters, a kernel size of 7 and dilations of [1, 2, 4, 8, 16]. After hyperparameter optimisation, the final TCN model architecture was trained using the combined training and validation set.

**Table S3**: Hyperparameter search space used for the grid search to define optimal parameters for the TCN model. Bold values indicate the final hyperparameters.

| **Hyperparameter** | **Values tested** |
|---|---|
| Number of filters | 16, **32**, 64 |
| Kernel Size | 3, 5, **7**, 10 |
| Dilations | [1,2,4,8], **[1,2,4,8,16]** |

[2] F. Chollet and others, “Keras.” [Online]. Available: https://keras.io/

[3] T. O'Malley *et al.*, “KerasTuner.” [Online]. Available: https://github.com/keras-team/keras-tuner

[4] P. Rémy, “Temporal Convolutional Networks for Keras.” [Online]. Available: https://github.com/philipperemy/keras-tcn
